# Histidine-rich glycoprotein is not associated with thrombosis in a UK Biobank Mendelian randomisation analysis

**DOI:** 10.64898/2026.07.20.26358305

**Authors:** Yuncheng Duan, Hamish M. Aitken-Buck, Peter MacCallum, Jeffrey I. Weitz, Ajay K. Kakkar, Andrew S. Allen

## Abstract

**Background:** Histidine-rich glycoprotein (HRG) is a regulator of coagulation that has been linked to experimental thrombosis, but its causal role in human thrombotic disease remains unclear.

**Objectives:** To determine whether HRG has a role in thrombosis, we evaluated the association between genetically determined HRG variation and thrombosis risk using Mendelian randomisation (MR).

**Methods:** We performed a two-sample MR analysis in UK Biobank using non-overlapping samples. Separate genome-wide association studies (GWAS) were conducted to identify single-nucleotide polymorphisms (SNPs) associated with circulating HRG protein levels and to estimate SNP associations with thrombosis outcomes. Genetic instruments were derived from the HRG GWAS measurements and applied to assess associations with overall, venous, and arterial thrombosis. Sensitivity analyses using multiple MR methods were undertaken, alongside adjusted logistic regression models in participants with measured HRG levels.

**Results:** Among 30,680 participants with HRG measurements, GWAS identified multiple loci associated with HRG levels, with the strongest signal at the rs9898 SNP (β=0.52; P=1.1×10^−306^). Single instrument MR found no association between HRG protein levels predicted by the rs9898 SNP and risk of overall thrombosis (β=−0.00660; P=0.622), venous thrombosis (β=0.0101; P=0.650) and arterial thrombosis (β=−0.0137; P=0.384). Similar null findings were observed using multi-instrument MR approaches. In complementary analyses, measured HRG levels were not associated with thrombosis after adjustment for age, sex, and C-reactive protein, and results were unchanged after stratification by rs9898 genotype.

**Conclusion:** Genetically determined variation in HRG is not associated with thrombotic risk, indicating that HRG-related coagulation phenotypes do not translate into clinically meaningful thrombosis.

## 1. INTRODUCTION

Thrombosis results from the convergence of multiple biological pathways that together promote a hypercoagulable state. Large-scale genome-wide association studies (GWAS) have identified numerous loci influencing the risk of venous and arterial thrombosis, including well-established variants in coagulation factor genes such as F5 and F11, as well as additional loci shared across thrombotic phenotypes [1–4]. These findings underscore the importance of genetically mediated perturbations in haemostatic balance as determinants of thrombotic risk.

Histidine-rich glycoprotein (HRG), encoded at chromosome 3q28–q29, is a liver-derived plasma protein that circulates at high concentrations and binds multiple ligands involved in coagulation and fibrinolysis, including factor XIIa, plasminogen, fibrinogen, heparin, and zinc ions [5–8]. Experimental manipulation of HRG levels *in vitro* and in animal models alters intrinsic pathway–dependent coagulation kinetics, reflected by changes in activated partial thromboplastin time (aPTT) without consistent effects on prothrombin time [7,9]. Circulating HRG concentrations are highly heritable, with genetic factors accounting for approximately 60–70% of inter-individual variation [10,11]. GWAS across multiple population-based cohorts have identified the HRG locus as the primary genetic determinant of plasma HRG levels, most prominently implicating the common missense variant rs9898, which has also been associated with aPTT [12–20].

Experimental and observational evidence has suggested a potential role for HRG in thrombosis. HRG-deficient mice exhibit enhanced thrombin generation and accelerated thrombus formation at sites of vascular injury in several experimental models [21–24]. Similarly, in humans, circulating HRG levels have been linked to clot structure and post-thrombotic syndrome, and rare HRG variants have been reported in individuals with venous thromboembolism (VTE) [25,26]. However, these observations do not establish causality and may be confounded by inflammatory or disease-related processes. Consistent with this uncertainty, direct association analyses of common HRG coding variants, including rs9898, have not demonstrated a clear relationship with venous thrombosis risk [27].

Mendelian randomisation (MR) offers a robust genetic framework to test causal relationships between modifiable exposures and disease outcomes, minimising confounding and reverse causation [28]. In thrombosis research, MR has distinguished non-causal biomarkers from traits with clear causal effects, including specific coagulation factors and platelet-related pathways [29–34]. Against this background, we used large-scale genetic and proteomic data from the UK Biobank [35] to clarify the relationship between genetically determined HRG levels and thrombosis. We identified robust genetic determinants of circulating HRG and applied two-sample MR to evaluate the causal relationship between genetically determined HRG protein levels and overall thrombosis, VTE, or arterial thrombosis, supported by complementary adjusted association analyses.

## 2. METHODS

### 2.1. Study Design

We performed a two-sample MR study to evaluate the relationship between genetically proxied HRG levels and the risk of thrombotic disease. Genetic instruments were identified from a GWAS of HRG protein levels measured in UK Biobank participants using UK Biobank Pharma Proteomics Project (UKB-PPP) data. The HRG and thrombosis GWAS samples were constructed to be non-overlapping: thrombotic disease cases were excluded from the HRG GWAS sample, and participants included in the HRG GWAS were excluded from the thrombosis control set. We prioritised rs9898 as the primary single-variant instrument because it represents a cis-HRG signal at the HRG locus. In addition, we performed multi-variant MR using five independent HRG-associated SNPs from the UKB-PPP analysis as instruments for circulating HRG levels. These instruments were used to assess associations with overall thrombosis, VTE, and arterial thrombosis outcomes in UK Biobank. Adjusted direct association analyses using logistic regression of measured HRG protein levels were conducted in parallel to support interpretation of the MR findings.

### 2.2. UK Biobank Participants, Genotyping, and Disease Phenotyping

This study utilised data from UK Biobank, a large prospective cohort including over 500,000 participants aged 40 to 69 years at recruitment [35]. Individuals were enrolled between 2006 and 2010 across assessment centres throughout England, Scotland, and Wales, with comprehensive genetic, phenotypic, and linked health record data available for analysis. Genotyping was performed on DNA extracted from blood samples collected at baseline assessment visits. Genotype data are available for 488,377 participants and were generated using two closely related Affymetrix arrays with largely overlapping marker content. Genotypes were centrally phased and imputed by UK Biobank using the Haplotype Reference Consortium and UK10K/1000 Genomes reference panels. In this study, we used the version 3 autosomal imputed genotype data in BGEN format for chromosomes 1-22, comprising approximately 93.1 million autosomal variants before analysis-specific quality control [36].

We defined thrombotic disease phenotypes by creating a curated phenotype table of International Classification of Disease (ICD)-9 and ICD-10 diagnosis codes (see ***Online Supplementary Information***). The disease phenotype definitions focused on VTE and arterial thrombotic events and incorporated multiple UK Biobank data sources, including hospital inpatient records, death registry data, self-reported diagnoses, and self-reported procedure fields. A previous study of the UK Biobank database found that no single data source (i.e. primary care records or participant self-reporting) sufficiently captures all cases and recommended using all available data sources [37]. Arterial thrombotic events encompassed myocardial infarction, non-cardioembolic ischaemic stroke, and peripheral vascular disease. For myocardial infarction, we also included algorithmically defined UK Biobank outcomes. Cases were defined as participants with evidence of thrombotic disease based on the curated diagnosis, procedure, and algorithmic outcome criteria. Exclusions based on ICD-9 or ICD-10 codes were applied to the case definitions of VTE and non-cardioembolic stroke, as detailed in the ***Online Supplementary Information***.

### 2.3. Ethics Approval

This study used data from UK Biobank under approved application number 194362. UK Biobank received ethical approval from the North West Multi-centre Research Ethics Committee (reference 11/NW/0274), and all participants provided written informed consent for participation and for the use of their data and samples in approved health-related research.

### 2.4. HRG Protein Measurement

HRG protein levels were determined using the Inflammation II panel of the Olink Explore 3072 plasma proteomics platform (UKB-PPP protein ID: HRG:P04196:OID30770:v1), which employs proximity extension assay (PEA) technology as previously described [17]. The UKB-PPP cohort comprised 54,219 participants selected from the wider UK Biobank population, including 46,595 randomly selected individuals, 6,376 participants selected by consortium members, and 1,268 participants from the COVID-19 imaging study. Of these, Olink Explore 3072 proteomic data were available for 48,893 participants, covering 2,942 proteins, with protein abundance reported as normalised protein expression (NPX) values on a log_2_ scale. HRG protein levels were available for 44,605 participants.

### 2.5. GWAS Analyses for HRG Protein Level and Thrombosis Outcomes

Genome-wide association analyses were conducted using UK Biobank version 3 autosomal imputed genotype data in BGEN format. Genotype quality control followed the same framework for the HRG protein GWAS and thrombosis GWAS. Briefly, genotype files were restricted to the analysis sample and converted to PLINK2 format. Standard quality control steps were applied, including filtering for variant- and sample-level missingness. Relatedness was estimated using the KING method in PLINK2, and one individual from each related pair was removed based on a kinship cutoff. Duplicate variants were removed, and variants were LD-pruned using a sliding-window approach (window size 50 variants, step size 5 variants, r^2^ threshold = 0.2). Principal component analysis was then performed on the resulting set of unrelated, LD-pruned genotypes. We restricted downstream analyses to the genetically defined Caucasian subset provided by UK Biobank (field 22006). Within this ancestry-restricted sample, additional genotype quality control was performed, including variant- and sample-level missingness filtering, Hardy–Weinberg equilibrium filtering, relatedness filtering, and principal component analysis. The final genetic principal components were retained as covariates in downstream association analyses.

A GWAS was conducted to identify variants associated with circulating HRG protein levels. The analysis was restricted to participants with measured plasma HRG levels after excluding individuals with thrombotic disease, yielding 37,645 individuals before genotype quality control and 30,679 individuals after quality control. Association testing was performed using SAIGE, with inverse-normalised HRG protein levels as the quantitative phenotype and sex and the top five genetic principal components included as covariates. Effect estimates from this analysis were used to define genetic instruments for subsequent MR analyses. A second GWAS was conducted using the same analytical framework to assess genetic associations with thrombotic outcomes. Before genotype quality control, the thrombosis GWAS phenotype comprised 74,905 cases, including 26,635 VTE and 48,270 arterial thrombosis cases, and 367,816 controls. After quality control, the final thrombosis GWAS sample included 58,216 cases, including 18,478 VTE and 39,738 arterial thrombosis cases, and 280,915 controls. Association testing was performed using SAIGE, with case-control status as the binary phenotype and sex and the top five genetic principal components included as covariates. Quantile–quantile plots and Manhattan plots were used to inspect the GWAS results.

### 2.6. Genetic Instrument Selection and MR Analysis

MR was undertaken to evaluate whether genetically determined variation in circulating HRG levels has a causal effect on thrombotic outcomes, including overall, arterial, and venous thrombosis. Genetic instrument variables were selected from SNPs identified in the HRG GWAS. The primary causal estimate was derived using the Wald ratio method. For multi-variant MR, we used five independent HRG-associated SNPs identified from the UKB-PPP HRG GWAS as instruments for circulating HRG levels. Multiple MR methods were used, including inverse-variance weighted, MR-Egger, weighted median, simple mode, and weighted mode.

### 2.7. Direct Association Analysis

To complement the MR analyses, we performed direct association analyses to assess the relationship between circulating HRG protein levels and thrombotic risk. These analyses were restricted to participants with available HRG measurements and were conducted using multivariable logistic regression. Separate models were fitted for any thrombosis, VTE, and arterial thrombosis. Each model included HRG protein level as the exposure and was adjusted for sex, birth year as a proxy for age, and CRP (UK Biobank field 30710) as a marker of systemic inflammation. We first analysed all rs9898 genotype groups together and then repeated the adjusted models after stratifying participants by rs9898 genotype (CC, CT, and TT).

## 3. RESULTS

### 3.1. GWAS Identification of SNPs Associated with HRG Protein Abundance and Thrombosis

The HRG protein and thrombosis GWAS analyses were conducted in independent UK Biobank samples. Principal component analysis defined genetically homogeneous subsets for each analysis (**Supplementary Figures 1** and **2**), and association testing was performed using SAIGE.

In the HRG GWAS, 30,679 participants remained after quality control. The quantile–quantile plot showed marked deviation from the null distribution, consistent with an excess of significant associations (**Supplementary Figure 3a**). The principal signals (**Figure 1a**) corresponded to previously reported HRG-associated loci [17], led by rs9898 on chromosome 3 (MAF = 0.329; β = 0.515; P = 1.10 × 10^−306^), followed by rs4934470 (chromosome 10; MAF = 0.273; β = −0.116; P = 8.55 × 10^−38^), rs1206245 (chromosome 1; MAF = 0.368; β = 0.0594; P = 9.75 × 10^−13^), rs1260326 (chromosome 2; MAF = 0.391; β = −0.0472; P = 1.07 × 10^−8^), and rs59950280 (chromosome 4; MAF = 0.334; β = 0.0430; P = 1.00 × 10^−6^). The variant rs1042445 was also significantly associated with HRG levels (MAF = 0.223; β = 0.0785; P = 4.79 × 10^−16^). These findings were consistent with prior large cohort studies [12,13,15–17], although the positive effect direction for rs9898 contrasts with the negative association reported by Underwood *et al*. in US and Irish cohorts [19].

**Figure 1.**
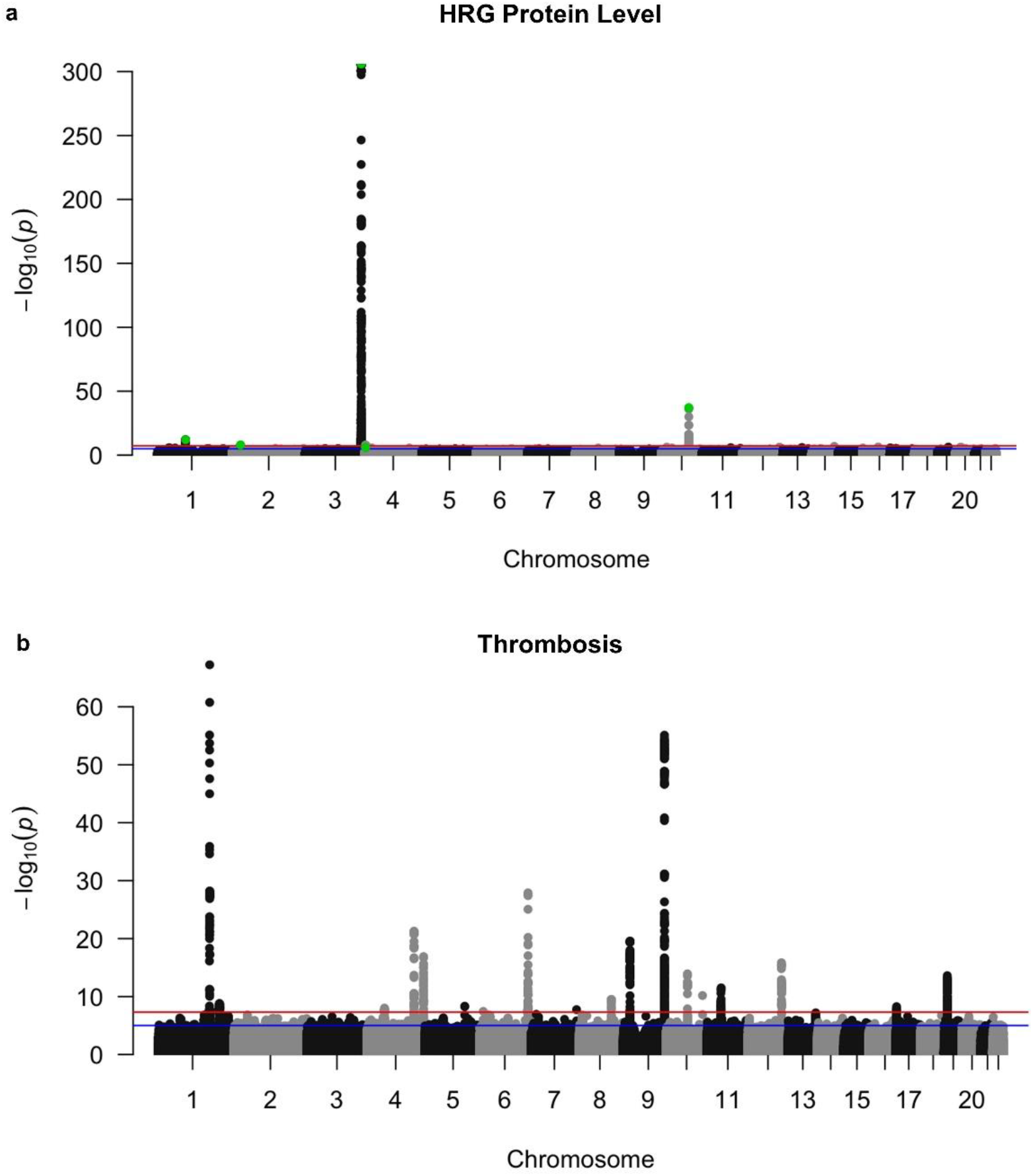
Genome-wide associations of plasma HRG levels and thrombosis in independent samples in UK Biobank. **a**, Manhattan plot for an association test identifying SNPs associated with HRG protein level in plasma. Protein levels were measured using the Olink PEA platform as part of the UKB-PPP. Highlighted in green are several HRG protein quantitative trait loci identified previously. **b**, Manhattan plot for an association test identifying genetic predictors of thrombosis. Association testing was conducted using SAIGE, with adjustment for sex and the top five genetic principal components. For **a**, N=30,679 participants. For **b**, N=58,216 cases and 280,915 controls. Abbreviations: GWAS = genome-wide association study; HRG = histidine-rich glycoprotein; PEA = proximity extension assay; SNP = single nucleotide polymorphism; UKB-PPP = UK Biobank Pharma Proteomics Project.

In the thrombosis GWAS, 58,216 cases and 280,915 controls remained after quality control. The any-thrombosis analysis identified genome-wide significant associations at several established thrombosis loci (**Figure 1b** and **Supplementary Figure 3b**), including variants at the *ABO, ZFPM2*, and *F2* loci [1,3]. We then examined HRG-associated variants of interest across all thrombosis phenotypes, including rs9898, the lead variant from the HRG GWAS, and rs1042445, previously reported by Underwood *et al*. [19]. Neither variant was significantly associated with overall thrombosis, VTE, or arterial thrombosis in single-variant GWAS analyses.

### 3.2. MR Analysis of HRG Protein Levels and Thrombosis Risk

Single-instrument MR for rs9898 using the Wald ratio method provided no evidence of an association between genetically determined HRG levels and overall thrombosis (**Table 1**). The corresponding estimates were also null for VTE (β = 0.0101; P = 0.650) and arterial thrombosis (β = −0.0137; P = 0.384). An analogous single-instrument MR analysis for rs1042445 likewise showed no association with overall thrombosis risk (β = −0.0236; P = 0.813).

**Table 1.** Estimates from Mendelian randomisation with single and multiple instruments for the effect of genetically determined HRG protein levels on overall thrombosis.

| Outcome = Any thrombosis<br>SNP | $\beta$ coefficient | P value |
| --- | --- | --- |
| **Single instrument MR (Wald ratio method)** |  |  |
| rs9898 | -0.00660 | 0.622 |
| **Multi-instrument MR** |  |  |
| MR Egger | -0.00634 | 0.863 |
| Weighted median | -0.00519 | 0.688 |
| Inverse variance weighted | 0.000273 | 0.990 |
| Simple mode | -0.0120 | 0.880 |
| Weighted mode | -0.00679 | 0.646 |

For the multi-instrument MR, the following SNPs identified in the HRG protein GWAS were used: rs9898, rs4934470, rs1206245, rs1260326, and rs59950280. Overall thrombosis encompasses VTE and arterial thrombosis, including myocardial infarction, non-cardioembolic ischaemic stroke, and peripheral vascular disease. Abbreviations: GWAS = genome-wide association study; HRG = histidine-rich glycoprotein; MR = Mendelian randomisation; SNP = single nucleotide polymorphism; VTE = venous thromboembolism.

Multi-instrument MR using the five HRG-associated variants identified in our study and reported in the original UKB-PPP study showed similar results [17]. Across MR-Egger, weighted median, inverse variance weighted, simple mode, and weighted mode methods, estimates were close to the null and P values did not indicate an association with overall thrombosis risk (**Table 1**).

### 3.3. Logistic Regression Analysis of HRG Levels with Thrombosis Risk

In multivariable logistic regression analyses, measured HRG protein levels were not associated with risk of any thrombosis (β = 0.0336; P = 0.473), VTE (β = 0.0827; P = 0.286), or arterial thrombosis (β = 0.0194; P = 0.724) (**Table 2**). Female sex and later birth year were associated with lower thrombosis risk, whereas higher CRP levels were associated with increased risk.

**Table 2.** Multivariable logistic regression analysis of overall thrombosis, venous thromboembolism, and arterial thrombosis risk.

| Outcome Variable | $\beta$ coefficient | P value |
| --- | --- | --- |
| **Overall thrombosis (n = 32,874)** |  |  |
| HRG protein | 0.0336 | 0.473 |
| Sex | -0.473 | $3.84 \times 10^{-54}$ |
| CRP | 0.0370 | $7.87 \times 10^{-37}$ |
| Birth year | -0.0719 | $8.10 \times 10^{-249}$ |
| **Venous thromboembolism (n = 29,069)** |  |  |
| HRG protein | 0.0827 | 0.286 |
| Sex | -0.124 | 0.0137 |
| CRP | 0.0348 | $1.53 \times 10^{-17}$ |
| Birth year | -0.0604 | $1.86 \times 10^{-67}$ |
| **Arterial thrombosis (n = 31,131)** |  |  |
| HRG protein | 0.0194 | 0.724 |
| Sex | -0.632 | $1.03 \times 10^{-68}$ |
| CRP | 0.0344 | $2.38 \times 10^{-26}$ |
| Birth year | -0.0776 | $2.46 \times 10^{-202}$ |

Overall thrombosis encompasses VTE and arterial thrombosis, including myocardial infarction, non-cardioembolic ischaemic stroke, and peripheral vascular disease. Abbreviations: CRP = C-reactive protein; HRG = histidine-rich glycoprotein; SD = standard deviation; VTE = venous thromboembolism.

Given the strong association between rs9898 genotype and HRG protein levels, we conducted stratified analyses to explore whether this missense variant might influence HRG function and thereby modify the relationship between HRG levels and thrombotic risk. Specifically, we repeated the multivariable regression analyses after stratifying participants by rs9898 genotype (CC, CT, and TT). Consistent with the primary analyses, HRG protein levels were not associated with risk of overall thrombosis, VTE, or arterial thrombosis within any genotype group (**Table 3**). These findings suggest that any functional effects of this variant do not translate into differential associations with clinical thrombotic outcomes.

**Table 3.** Multivariable logistic regression results of any thrombosis, venous thromboembolism, and arterial thrombosis by rs9898 variant genotype.

| Outcome<br>rs9898 genotype (No. of cases)<br>Variable | $\beta$ coefficient | P value |
| --- | --- | --- |
| **Overall thrombosis** |  |  |
| CC (n = 14,672) |  |  |
| HRG protein | 0.0560 | 0.446 |
| Female sex | -0.483 | $4.42 \times 10^{-26}$ |
| CRP | 0.0345 | $3.41 \times 10^{-16}$ |
| Birth year | -0.0704 | $4.24 \times 10^{-107}$ |
| CT (n = 14,287) |  |  |
| HRG protein | 0.0214 | 0.778 |
| Female sex | -0.475 | $1.50 \times 10^{-24}$ |
| CRP | 0.0417 | $6.07 \times 10^{-20}$ |
| Birth year | -0.0748 | $3.71 \times 10^{-116}$ |
| TT (n = 3,468) |  |  |
| HRG protein | 0.0471 | 0.764 |
| Female sex | -0.449 | $1.98 \times 10^{-6}$ |
| CRP | 0.0286 | $2.53 \times 10^{-3}$ |
| Birth year | -0.0681 | $2.95 \times 10^{-26}$ |
| **Venous thromboembolism** |  |  |
| CC (n = 12,956) |  |  |
| *HRG protein* | 0.0481 | 0.696 |
| *Female sex* | -0.101 | 0.183 |
| *CRP* | 0.0339 | $6.66 \times 10^{-9}$ |
| *Birth year* | -0.0513 | $3.44 \times 10^{-23}$ |
| CT (n = 12,635) |  |  |
| *HRG protein* | 0.0120 | 0.924 |
| *Female sex* | -0.145 | 0.0573 |
| *CRP* | 0.0311 | $3.60 \times 10^{-6}$ |
| *Birth year* | -0.0671 | $6.39 \times 10^{-36}$ |
| TT (n = 3,085) |  |  |
| *HRG protein* | 0.457 | 0.07 |
| *Female sex* | -0.167 | 0.279 |
| *CRP* | 0.0524 | $1.11 \times 10^{-5}$ |
| *Birth year* | -0.0738 | $1.22 \times 10^{-11}$ |
| **Arterial thrombosis** |  |  |
| CC (n = 13,909) |  |  |
| *HRG protein* | 0.0712 | 0.407 |
| *Female sex* | -0.654 | $9.51 \times 10^{-34}$ |
| *CRP* | 0.0305 | $8.28 \times 10^{-11}$ |
| *Birth year* | -0.0796 | $2.41 \times 10^{-94}$ |
| CT (n = 13,530) |  |  |
| *HRG protein* | 0.0266 | 0.766 |
| *Female sex* | -0.626 | $3.37 \times 10^{-30}$ |
| *CRP* | 0.0433 | $3.68 \times 10^{-18}$ |
| *Birth year* | -0.0790 | $2.49 \times 10^{-91}$ |
| TT (n = 3,279) |  |  |
| *HRG protein* | -0.139 | 0.451 |
| *Female sex* | -0.584 | $1.99 \times 10^{-7}$ |
| *CRP* | 0.00902 | 0.483 |
| *Birth year* | -0.0655 | $4.06 \times 10^{-18}$ |

Overall thrombosis encompasses VTE and arterial thrombosis, including myocardial infarction, non-cardioembolic ischaemic stroke, and peripheral vascular disease. Abbreviations: CRP = C-reactive protein; HRG = histidine-rich glycoprotein; VTE = venous thromboembolism.

## 4. DISCUSSION

We assessed the relationship between genetically determined circulating HRG protein levels and the risk of thrombosis. Among more than 30,000 UK Biobank participants with measured HRG levels in the UKB-PPP platform, we identified five SNPs significantly associated with HRG protein abundance, with rs9898 emerging as the primary genetic determinant. Despite these strong genetic associations, and despite the established roles of HRG in coagulation and thrombosis in experimental models, we found no evidence that genetically determined HRG protein levels were associated with thrombosis in MR or adjusted regression analyses. These findings reveal a possible dissociation between HRG-associated molecular or laboratory phenotypes reported in prior work and clinical thrombotic outcomes, motivating closer examination of how HRG variation, particularly post-translational modification, may influence protein biology without necessarily translating into disease risk.

Our GWAS of UK Biobank participants with available genetic and proteomic data identified several loci associated with circulating HRG levels measured using the Olink platform, with the strongest signal observed for the minor T allele of rs9898. This variant showed a robust positive association with measured HRG abundance, consistent with independent reports using Olink-based platforms, including analyses in the UK Biobank and the EPIC Norfolk cohort [12,15,17]. Concordant findings have also been demonstrated using mass spectrometry-based proteomics, which provides greater specificity through direct quantification of peptides. In a recent study, rs9898 was strongly associated with increased HRG levels in both discovery and replication cohorts, providing orthogonal validation of this association [16]. Similar positive associations have also been reported in Icelandic populations using both Olink and SomaScan platforms, although modest cross-platform correlations suggest that different technologies capture overlapping but not identical aspects of HRG biology [12,13].

In contrast, a recent study reported a negative association between rs9898 and HRG levels measured using a custom antibody-based AlphaLISA assay designed to estimate absolute protein concentrations [19]. Thus, although rs9898 shows a reproducible association with HRG levels across studies, the direction of effect appears to depend on the proteomic platform used. A plausible explanation for these opposing results lies in methodological differences among protein quantification assays, as well as the biological consequences of the rs9898 missense variant. The rs9898 T allele results in a proline-to-serine substitution at position 204 of HRG [36], creating a consensus glycosylation motif spanning amino acids 202–204 and introducing a novel glycosylation site at Asn202 [37]. In affinity-based proteomic assays such as Olink or SomaScan, measured signal intensity does not necessarily reflect true protein abundance, because post-translational modification, altered protein conformation, or amino acid substitutions within or near antibody-binding epitopes can affect target–binder interactions and detection independently of circulating concentration [38]. Further research is required to determine the direct effects of individual SNPs on HRG antibody affinity and, in turn, protein quantification.

Using the coding variants linked to HRG protein levels in our GWAS as genetic instruments, we evaluated whether HRG-associated variation causally influences thrombotic risk in the UK Biobank. Across single- and multi-instrument MR analyses, we found no evidence that SNPs predictive of HRG protein levels were associated with thrombosis, whether considered overall or stratified by VTE and arterial thrombosis outcomes. HRG protein levels were also not associated with the risk of thrombosis in adjusted regression analyses. Overall, our results are consistent with findings from the MARTHA study, which reported no association between rs9898 and venous thrombosis in more than 1,500 cases and controls [27]. Although that study was likely underpowered to detect modest effects, the absence of association aligns with our observations, collectively suggesting that genetically determined differences in HRG protein levels linked to common variants do not translate into a materially altered risk of clinical thrombotic disease.

This null result contrasts with substantial MR evidence supporting causal roles for several core haemostatic traits in thrombotic disease. Large two-sample MR studies have demonstrated causal associations between genetically determined increases in intrinsic pathway components, most notably factor XI, and higher platelet counts, which are associated with greater risk of ischaemic stroke [29,30]. Broader MR analyses have further shown that genetically proxied inhibition of fibrinogen and factors II and XI is associated with reduced risks of VTE and ischaemic stroke [31,33]. In this context, the absence of a causal signal for genetically determined HRG variation suggests that, unlike these central haemostatic components, HRG is unlikely to be a primary determinant of thrombotic risk in humans, despite its putative modulatory role in coagulation.

This interpretation also appears to conflict with experimental data supporting HRG as a negative regulator of coagulation. HRG-deficient mice exhibit enhanced thrombin generation, shortened clotting times, and accelerated thrombus formation in arterial injury and polyphosphate-induced thrombosis models, phenotypes that are attenuated by HRG supplementation [21–24]. However, these models reflect complete or near-complete loss of HRG function under strongly procoagulant conditions, whereas single HRG variants, such as rs9898, produce more subtle structural changes. Accordingly, isoform-level variation associated with specific HRG SNPs is unlikely to recapitulate these pronounced experimental phenotypes or substantially influence clinical thrombosis risk.

Our study has limitations. First, HRG protein levels were measured using the Olink proteomics platform, which provides relative rather than absolute quantification and reflects epitope-detectable abundance. Although well-suited to large-scale proteomics, this affinity-based method may be affected by protein structure or post-translational modifications, such as glycosylation [38]. Second, our analyses were restricted to individuals of European ancestry, limiting generalisability to populations in whom HRG levels, allele frequencies, and baseline thrombosis risk may differ; however, prior work suggests consistent rs9898–aPTT associations across European American and African American groups [20]. Third, VTE events could not be reliably classified as provoked or unprovoked, which may dilute associations if HRG primarily affects idiopathic disease. In addition, VTE case definitions relied on UK Biobank data sources, which have poor concordance, as most events are captured in only one source with minimal overlap [39]. This heterogeneity in ascertainment may introduce misclassification, bias, and sociodemographic differences between cases. Finally, MR estimates reflect lifelong genetically determined differences in HRG rather than acute or disease-driven changes and therefore may not directly predict the effects of therapeutic modulation.

In conclusion, we show that several coding variants associated with the HRG gene, most prominently rs9898, predict variation in circulating HRG protein levels. However, in both MR and adjusted observational analyses, neither genetically determined differences linked to HRG coding variants nor measured HRG protein levels were associated with clinical thrombotic outcomes. These findings suggest that common structural variation in HRG can influence protein abundance and laboratory phenotypes without translating into clinically meaningful thrombosis risk, supporting a modulatory rather than causal role for HRG in human thrombosis.

## AUTHOR CONTRIBUTIONS

Y. Duan, H.M. Aitken-Buck, P. MacCallum, J.I. Weitz, A.K. Kakkar, and A.S. Allen all contributed to the concept, design, and conduct of the study. Y. Duan, and A.S. Allen conducted the statistical analysis. Y. Duan, H.M. Aitken-Buck, P. MacCallum, J.I. Weitz, and A.S. Allen drafted the manuscript. All authors contributed to result interpretation. A.K. Kakkar, and A.S. Allen handled funding and supervised the study. All authors approved the final version of the manuscript.

## ACKNOWLEDGEMENTS

Statistical and administrative support was provided by Karen Pieper (Thrombosis Research Institute, UK). Statistical support was provided by Dr Meg E. Fluharty (formerly Thrombosis Research Institute, London, UK).

## DATA AVAILABILITY

UK Biobank data are available in accordance with their published data access procedures described at http://www.ukbiobank.ac.uk/using-the-resource/. Study specific information can be shared upon reasonable request and receipt of an analysis plan to Andrew S. Allen.

