## Supplementary Information for "Histidine-rich glycoprotein is not associated with thrombosis in a UK Biobank Mendelian randomisation analysis"

### Supplementary Figure 1

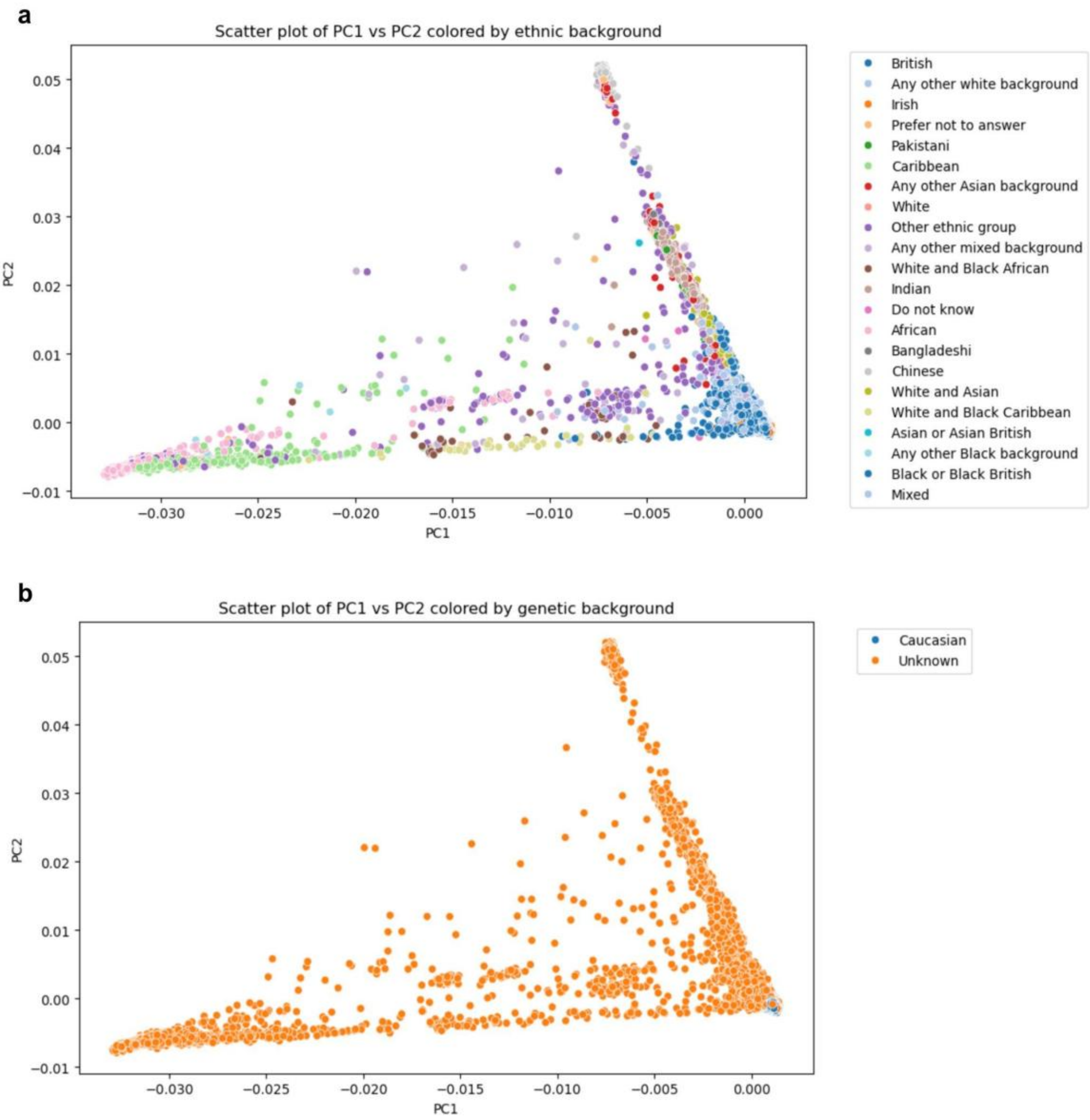


**Supplementary Figure 1. Principal component analysis of genetic ancestry in UK Biobank participants for the HRG GWAS.** The scatter plot in panel **a** shows the first two principal components (PC1 and PC2) derived from genome-wide genotype data, coloured by self-reported ethnic background, illustrating population structure within the cohort. Distinct clustering of participants reflects underlying genetic ancestry. Panel **b** shows the same principal component space coloured according to genetic ancestry classification after quality control filtering. Individuals within the defined European ancestry cluster, labelled as Caucasian, were retained for downstream analyses, while samples outside this cluster were excluded to minimise population stratification. Abbreviations: GWAS = genome-wide association study; HRG = histidine-rich glycoprotein.

#### Supplementary Figure 2

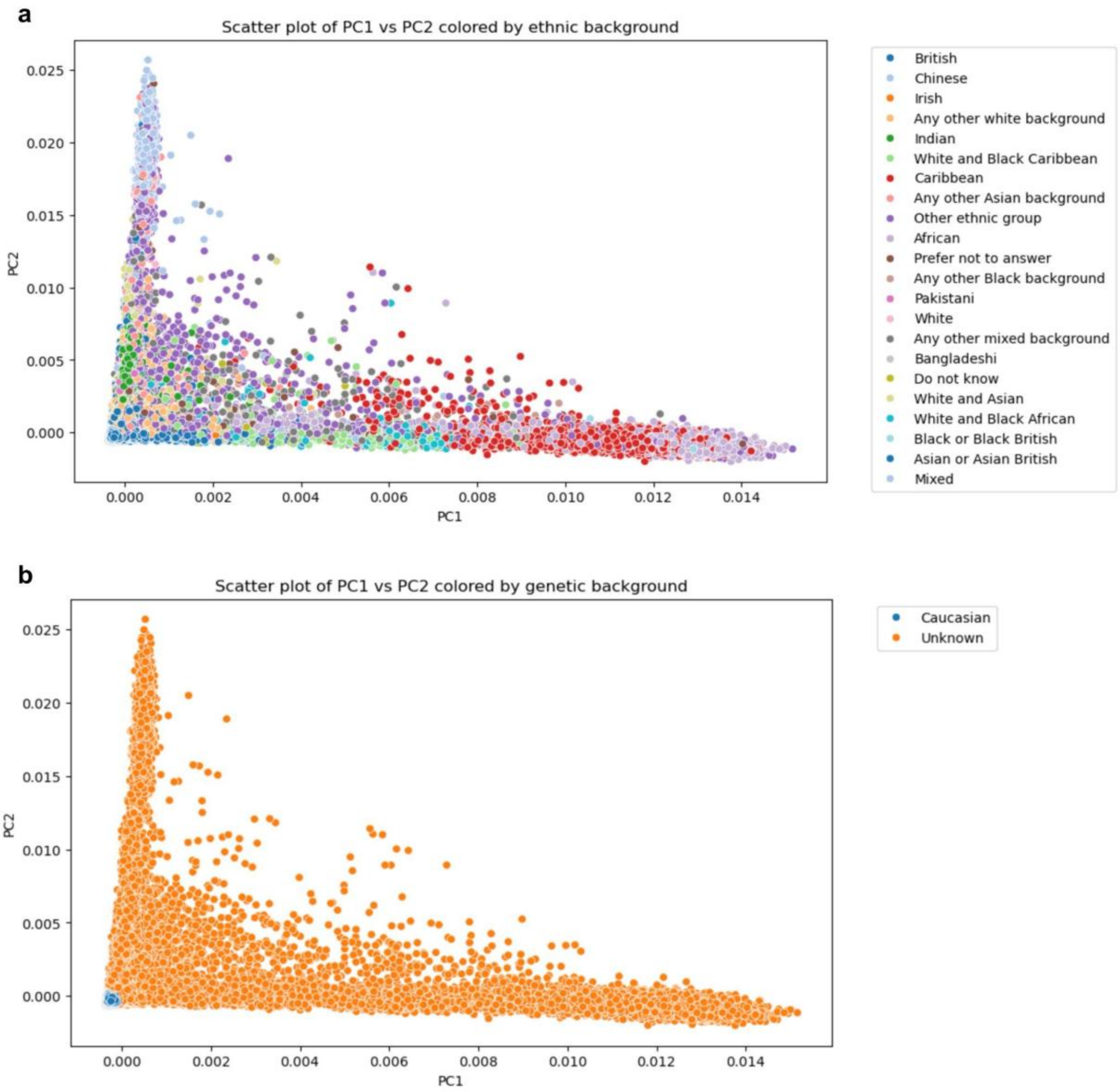


**Supplementary Figure 2. Principal component analysis of genetic ancestry for the thrombosis GWAS dataset.** The scatter plot in panel **a** shows the first two principal components (PC1 and PC2) derived from genome-wide genotype data, coloured by self-reported ethnic background, demonstrating population structure within the thrombosis GWAS cohort. Distinct clustering reflects underlying genetic ancestry across individuals. Panel **b** shows the same principal component space coloured by genetic ancestry classification following quality control. Individuals clustering within the European ancestry group, labelled as Caucasian, were retained for analysis, while samples outside this cluster were excluded to minimise population stratification and ensure a genetically homogeneous dataset for downstream GWAS and MR analyses. Abbreviations: GWAS = genome-wide association study; MR = Mendelian randomisation.

#### Supplementary Figure 3

**a**

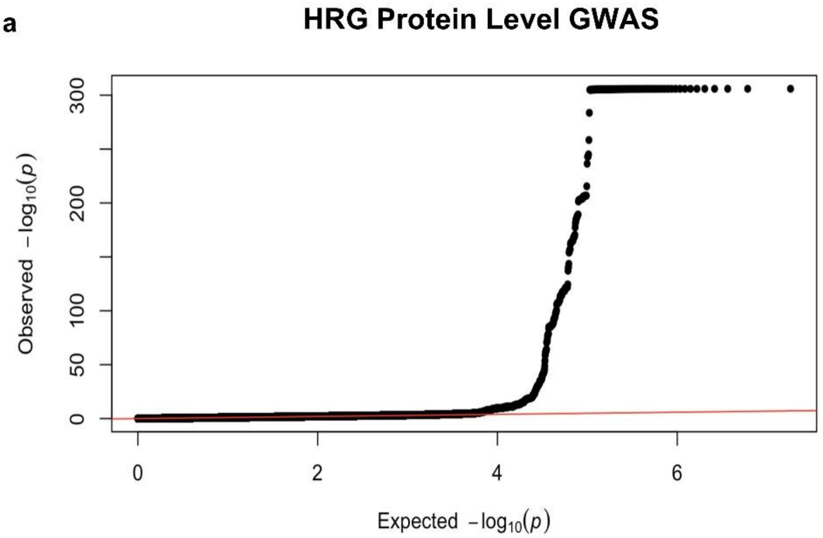


**b**

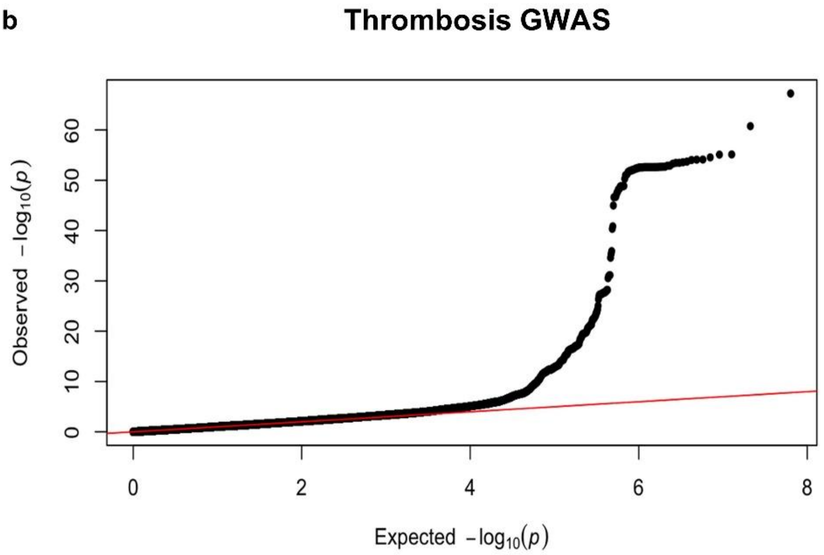


**Supplementary Figure 3. Quantile–quantile plots from the GWAS analyses of HRG protein level and thrombosis risk in UK Biobank.** Quantile–quantile plots for association tests identifying instruments for HRG protein level in plasma and thrombosis risk are shown in panels **a** and **b**, respectively. In both, there is marked deviation from the null distribution, indicating an excess of significant genetic associations. For **a**, N=30,679 participants. For **b**, N=58,216 cases and 280,915 controls. Abbreviations: GWAS = genome-wide association study; HRG = histidine-rich glycoprotein.
